# Efficacy and Safety of Glibenclamide in Patients with Post-Ischemic Stroke Cerebral Edema: A Systematic Review and Meta-Analysis

**DOI:** 10.1101/2025.06.04.25329003

**Authors:** Abdallah Khatatbeh, Basma AbuMahfouz, Sophia Braund, Ala’ Alrawashdeh, Neha Momin, Ahmad Al-Tanjy, Mohammad El-Ghanem

## Abstract

**Background:** The potential of Glibenclamide to improve functional outcomes in patients with acute ischemic stroke remains controversial. This study aims to evaluate the benefit of Glibenclamide for patients with acute ischemic stroke and at risk of malignant brain edema.

**Methods:** A comprehensive search was undertaken in January 2025 across several electronic databases, including PubMed, Scopus, Cochrane Library, Web of Science, and Embase. RCTs and observational studies were both considered eligible. Data extraction and analysis were done using Review Manager (RevMan) version 5.4.

**Results:** Six studies (four RCTs and two cohort studies) involving 1,244 patients were included; five were eligible for meta-analysis. Glibenclamide significantly reduced MMP-9 levels (MD = – 20.35; 95% CI: –23.65 to –17.04; P < 0.00001; I² = 0%) and midline shift (MD = –2.37 mm; 95% CI: –4.00 to –0.73; P = 0.005; I² = 83%). However, it did not significantly improve modified Rankin Scale scores or reduce the need for decompressive craniotomy. Glibenclamide was associated with a higher risk of hypoglycemia (RR = 3.30; 95% CI: 1.26–8.66; P = 0.02; I² = 0%) and serious adverse events (RR = 1.12; 95% CI: 1.01–1.24; P = 0.04; I² = 0%). No significant differences were observed in other adverse effects.

**Conclusion:** Glibenclamide may reduce inflammation and cerebral edema, but it increases the risk of hypoglycemia and serious adverse events. Given limitations in sample size and study variability, larger high-quality RCTs are needed to confirm efficacy and safety.

## 1. Introduction

Annually, 15 million people worldwide suffer from strokes (1). Strokes are the world’s second biggest cause of death, killing around 5.5 million individuals each year. The burden of stroke is shown by its high death and morbidity rates. Data also reveals that up to 50% of survivors develop chronic disabilities. Based on this, strokes are a major public health concern with considerable economic and social consequences. The public health burden of stroke is expected to increase in the coming decades as populations change demographically (2).

Cerebral edema is a potentially devastating consequence of ischemic stroke, and it is associated with high mortality rates and poor functional outcomes (3). It raises intracranial pressure, causes fast deterioration of neurological symptoms, and results in the formation of a cerebral hernia, making it a significant risk factor for poor outcomes after stroke. To date, the precise mechanism of cerebral edema after stroke is unknown. This limits progress in preventative and therapy measures, as well as drug development (4). Current guidelines for managing cerebral edema after an ischemic stroke are primarily supportive and limited, including the use of corticosteroids, osmotherapy, and surgical decompression (5).

Glibenclamide’s mode of action involves the blockage of ATP-sensitive K+ channels, which causes cell depolarization and insulin production. The drug’s extra pancreatic effect on the heart muscle and smooth muscle sites uses the same mechanism (6).

Based on a phase 3 double blind randomized trial published in 2024 by Sheth et al., intravenous Glibenclamide did not improve functional outcomes for patients with massive hemispheric infarction. Because the trial was terminated early, it lacked the necessary capacity to reach significant results. Future research is needed to determine the potential benefit of intravenous Glibenclamide in patients suffering from ischemic strokes (7). This systematic review and meta-analysis aims to evaluate and assess the efficacy and safety of Glibenclamide in patients with post-ischemic stroke cerebral edema. This review seeks to understand the role of Glibenclamide in decreasing mortality and morbidity in post-ischemic stroke patients.

## 2. Methodology

This meta-analysis was registered in the International Prospective Register of Systematic Reviews (PROSPERO) under the ID number of CRD420251032530. The study was conducted following the Preferred Reporting Items for Systematic Reviews and Meta-Analyses (PRISMA) guidelines and the Cochrane Handbook (8) (9).

### 2.1 Literature Search

On December 15, 2024, we performed a comprehensive search using PubMed, Cochrane, Scopus, Embase, and Web of Science databases. The search strategy included controlled vocabulary (e.g., MeSH) and free-text terms, using Boolean operators to combine concepts.

The search strategy included keywords such as (“Glibenclamide” OR “Glyburide” OR “Glibenclamida”) AND (“Stroke” OR “Ischemic Stroke” OR “Cerebrovascular Accident” OR “CVA” OR “Brain Infarction”) AND (“Cerebral Edema” OR “Brain Edema” OR “Brain Swelling” OR “Post-Stroke Edema” OR “Post-Infarction Edema” OR “Brain Edema”). No date or geographical restrictions were applied.

### 2.2 Eligibility Criteria and Study Selection

Eligible studies included randomized controlled trials (RCTs) and observational studies with the following PICOS eligibility criteria:

● **Population (P):** Adults with acute ischemic stroke complicated by cerebral edema.
● **Intervention (I):** Studies assessing the use of Glibenclamide or Glyburide as the intervention.
● **Comparator (C):** Comparator groups receiving standard therapy or placebo.
● **Outcomes (O):** Outcome measures, including efficacy (Modified Rankin Scale, midline shift), mortality, adverse events, and other relevant outcomes, reported in at least two studies.
● **Study Design (S):** Randomized controlled trials (RCTs), crossover trials, and observational studies.

Rayyan AI was used to organize studies across all databases and eliminate duplicate records

(10). Two authors independently evaluated titles, abstracts, and keywords for eligibility. Eligible studies underwent full-text evaluation. If the full text was unavailable, we attempted to contact the authors via email for more information; otherwise, the study was excluded. Discrepancies were addressed by consensus or by consulting with a third reviewer. We excluded observational studies, case reports, case series, conference abstracts, unpublished articles, inaccessible full text despite attempts to obtain additional information from authors, studies not published in English, animal studies, and research published only as abstracts or conference proceedings.

### 2.3 Quality Assessment

We used the Revised Cochrane risk-of-bias tool for randomized controlled trials (RoB-2) to assess the risk of bias in the clinical trials included (11). This comprehensive assessment addressed key domains such as randomization, sequence allocation concealment, deviations from intended interventions, and the use of appropriate statistical methods to estimate the effects of interventions, as well as outcome measurement, reporting biases, and overall bias risk. The methodological quality of the studies was evaluated as low risk, moderate concerns, or high risk of bias. The senior author’s ultimate decision resolved any disagreements. The Newcastle-Ottawa Scale was used for observational research (12). This tool allows investigators to use a point-based system to rate research as ‘good,’ ‘fair,’ or ‘bad.’ Any differences were settled through conversation, and a third author was consulted as needed.

### 2.4 Data Extraction

Two authors used a standardized method to extract data, systematically recording relevant information in a predefined Excel sheet. The sheet was constructed using a pre-specified uniform data extraction sheet that included the following domains: 1) characteristics of the studies included, 2) characteristics of the population of studies included, 3) risk of bias domains, and 4) outcome measures.

Study type, country, study duration, total number of patients, main outcomes, Glibenclamide dose, inclusion criteria, and conclusions were in the first domain. Patient demographics were captured in the second domain, including age, gender, comorbidities such as hypertension, diabetes mellitus, atrial fibrillation, transient ischemic stroke, and stroke etiology. Finally, the third domain included study outcomes, safety outcomes (Modified Rankin Scale (mRS) and midline shift), mortality, adverse events, and other relevant outcomes reported in at least two studies. Any discrepancies were resolved through consensus or by consulting the senior author.

### 2.4 Calculation of Missing Data

When data was reported as medians with interquartile ranges (IQR), it was converted to means and standard deviations (SD) using formulas by Wan et al. (13). If SD was not reported, it was derived from the standard error (SE) using SD = SE × √n (14). When the mean change (MC) between baseline and endpoint was missing, it was calculated as MC = M_post – M_pre. If the SD of the MC was unavailable, it was estimated using SD = √ (SD²_pre + SD²_post) (15). Missing effect sizes (d) were calculated as d = (M_treatment – M_control) / SD_pooled, where SD_pooled = √. Effect size correlation (r) was computed using r = d / √ (d² + 4).

### 2.5 Statistical Analysis

The standardized mean difference (SMD) was employed as the primary outcome measure. A random-effects model was applied to account for variability across studies. Between-study heterogeneity (τ²) was estimated using Hedges’ method. In addition to τ², heterogeneity was assessed using Cochran’s Q test and the I² statistic. When heterogeneity was present (i.e., τ² > 0), a prediction interval for the true effect size distribution was also reported. To identify potential outliers and influential studies, studentized residuals and Cook’s distances were examined. Studies with studentized residuals exceeding the 100 × (1 – 0.05 / [2k]) percentile of the standard normal distribution based on a Bonferroni-adjusted two-sided alpha of 0.05 for k studies were classified as potential outliers. Studies were considered influential if their Cook’s distance exceeded the median plus six times the interquartile range of all Cook’s distances.

### 2.6 Strategy for Data Synthesis

Statistical analyses were performed using the Comprehensive Meta-Analysis software. The standardized mean difference (SMD), along with its variance and standard error (SE), was calculated based on post-intervention values comparing treatment and control groups. For outcomes reported as medians with interquartile ranges, values were converted to means and standard deviations using the method proposed by Wan et al. All extracted data were subsequently pooled using the ‘meta’ package in R. Meta-analysis results were presented through forest plots and aggregated forest plots. A fixed-effect model (Mantel–Haenszel method) was used in the absence of significant heterogeneity; otherwise, a random-effects model (DerSimonian and Laird method) was applied. Statistical significance was defined as a p-value < 0.05. Heterogeneity was assessed using Cochran’s Q test and the I² statistic, with I² > 50% or p < 0.05 indicating substantial heterogeneity. Publication bias was evaluated using Egger’s regression test.

### 2.7 Publication Bias Assessment

Publication bias was not assessed, and a funnel plot was not done as the number of studies is less than 10.

## 3. Results

### 3.1 Literature Search Results

Through our preliminary search across multiple databases, we identified 2768 records with 1341 remaining after duplicate removal. Thirty-four records were screened for adherence to our inclusion and exclusion criteria. Ultimately, six studies, four randomized controlled trials, and two cohorts, were included in our meta-analysis. The PRISMA flow diagram, as shown in **Figure 1**, delineates this process.

**Figure 1:**
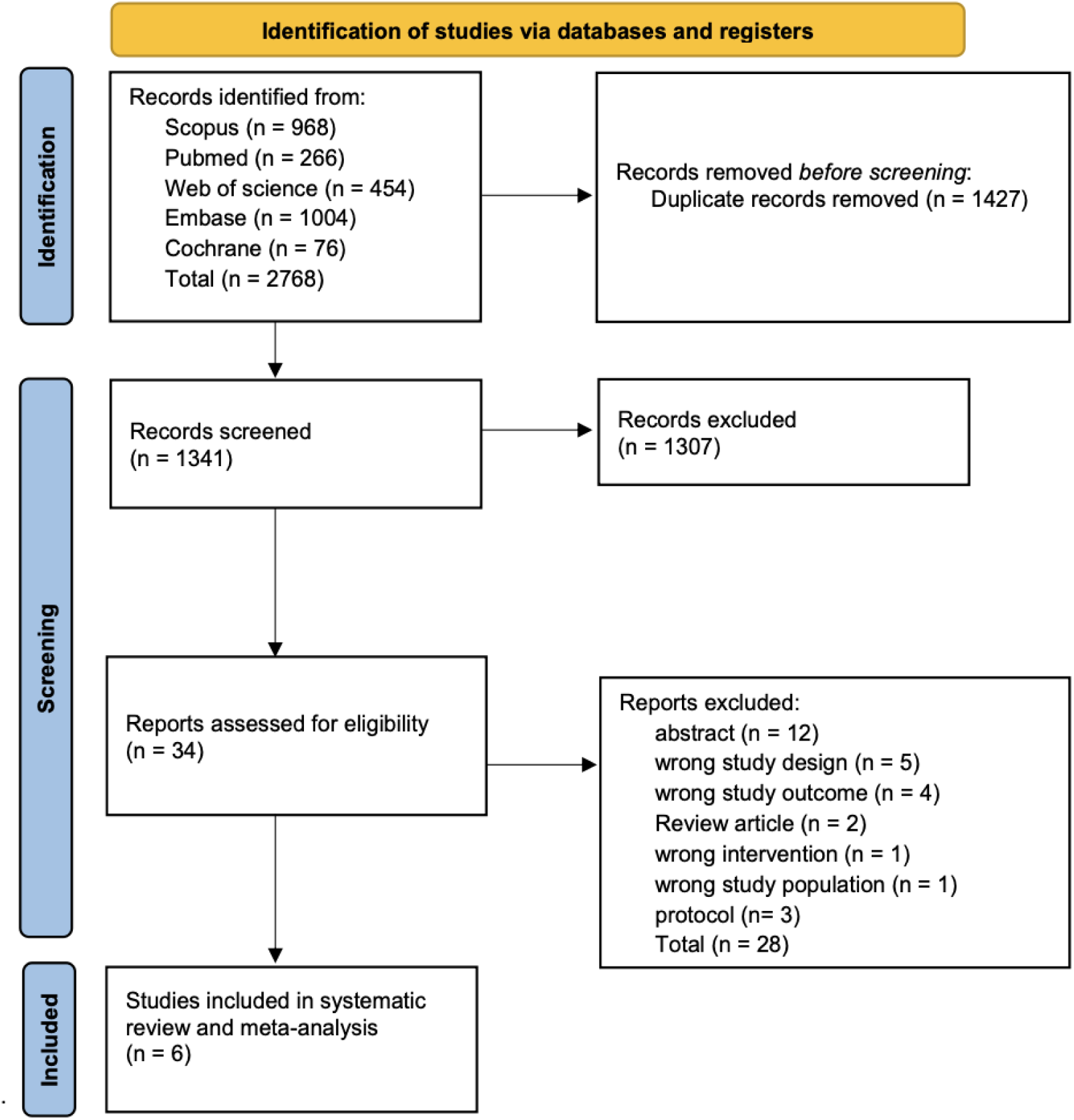
PRISMA Flow Diagram.

### 3.2 Characteristics of Individual Studies

A total of six studies with 1244 patients met the eligibility criteria and were included and a Glibenclamide dose ranging from 3 mg to 8.6 mg across varying time frames, 8.6 mg in most os studies. **Tables 1 and 2** include a summary of each of the included studies and their baseline characteristics.

**Table 1:** Summary of the Included Studies.

| Study ID | Country | Study design | Study period | Sample size, n | Glibenclamide Dose | Criteria of recruitment | Conclusion |
| --- | --- | --- | --- | --- | --- | --- | --- |
| **Vigilante 2023** | USA | Retrospective observational cohort | 10/2019 - 08/2021 | 95 | (2.5mg per nasogastric tube twice daily for 3 days) | Large hemisphere infarction + meeting the criteria for the CHARM trial. Key inclusion criteria for CHARM trials:<br>Age: $\geq 18$ years old<br>Heart Failure Severity: NYHA functional class II-IV<br>Left Ventricular Ejection Fraction (LVEF): $\leq 40\%$<br>ACE Inhibitor Status:<br>CHARM-Added: Currently taking a stable dose of an ACE inhibitor<br>CHARM-Alternative: Intolerant to ACE inhibitors | Glyburide was found to have a low risk of hypoglycemic events. Despite the small sample size, there was no signal of better clinical or imaging outcomes in treated patients. |
| **Sheth 2013** | USA | Prospective, open-label, multicenter, phase IIa pilot clinical trial | 02/2011 - 05/2012 | 10 | 3 mg/day | Inclusion criteria were baseline diffusion-weighted image lesion volume, 82 to 210 cm<sup>3</sup>; age 18 to 80 years; time from symptom onset to start of study drug infusion | The drug led to reduced hemispheric volume, hemorrhagic transformation, lesion growth, mortality, and improved neurological outcomes. |
|  |  |  |  |  |  | ≤10 hours. |  |
| **Sheth**<br>**2024** | Multiple stroke centers in 21 countries | Phase 3: double-blind placebo-controlled randomized trial | 08/2018 - 05/2023 | 535 | 8.6 mg over 72 h | Patients who had a large stroke (either by an ASPECTS of 1–5 or by an ischemic core lesion volume of 80–300 mL on CT perfusion or MRI diffusion-weighted imaging). Individuals within the age group of 18-85 years old with an acute ischemic stroke within 10 hours of stroke onset. NIHSS score of 10 or higher, absence of significant premorbid disability. | Results did not show improvement in functional outcome at 90 days in patients treated with Glibenclamide. |
| **Sheth**<br>**2016** | USA | Phase 2 Double-blind RCT | 05/2013 - 04/2015 | 86 | 8.6 mg over 72 h | Participants were aged 18–80 years and had a clinical diagnosis of large anterior circulation hemispheric infarction for less than 10 h from the time last known to be neurologically healthy, confirmed by a baseline diffusion weighted image (DWI) lesion volume of 82–300 cm<sup>3</sup>. Participants undergoing an endovascular thrombectomy were not eligible. | The study did not demonstrate that intravenous Glibenclamide would reduce the need for decompressive craniectomy. However it showed intravenous Glibenclamide is safe in critically ill patients after an ischemic stroke and there was non-significant reduction in mortality and reduced brain swelling. |
| Huang<br>2019 | China | Retrospective<br>-prospective<br>cohort study | 01/ 2017 -<br>08/2017 | 213 | 1.25 mg loading<br>dose, followed by<br>0.625 mg every 8<br>hours for 5 days | Patients were recruited if they<br>(a) aged between 18 and 80<br>years; (b) abled to receive<br>study drug within 10 hours of<br>the stroke onset; (c)<br>presented with an acute<br>anterior ischemic stroke and<br>an National Institute of Health<br>Stroke Score (NIHSS) of $\geq 8$ (d)<br>had a prestroke modified<br>Rankin Scale (mRS) of $\leq 1$ (e)<br>had not been on GBC or other<br>sulfonylureas within one<br>month prior to stroke; and (f)<br>did not suffered from other<br>end-stage malignant diseases<br>that might confound<br>prognosis. | Low-dose oral Glibenclamide is<br>safe and might reduce brain<br>edema and severe disability.<br>Functional outcomes were not<br>significantly improved, possibly<br>due to the small sample size<br>and study limitations. |
| Huang<br>2023 | China | Randomized,<br>double-blind,<br>placebo-controlled<br>trial | 01/2018 -<br>05/2022 | 305 | 1.25 mg loading<br>dose, followed by<br>0.625 mg every 8<br>hours for 5 days | Acute ischemic stroke treated<br>with rtPA within 4.5 hours of<br>symptom onset, no severe<br>heart disease, blood glucose<br>$\geq 3.0$ mmol/L | Glibenclamide did not improve<br>functional outcomes at 90 days<br>but was safe with reduced<br>MMP-9 levels. |

**Table 2:**
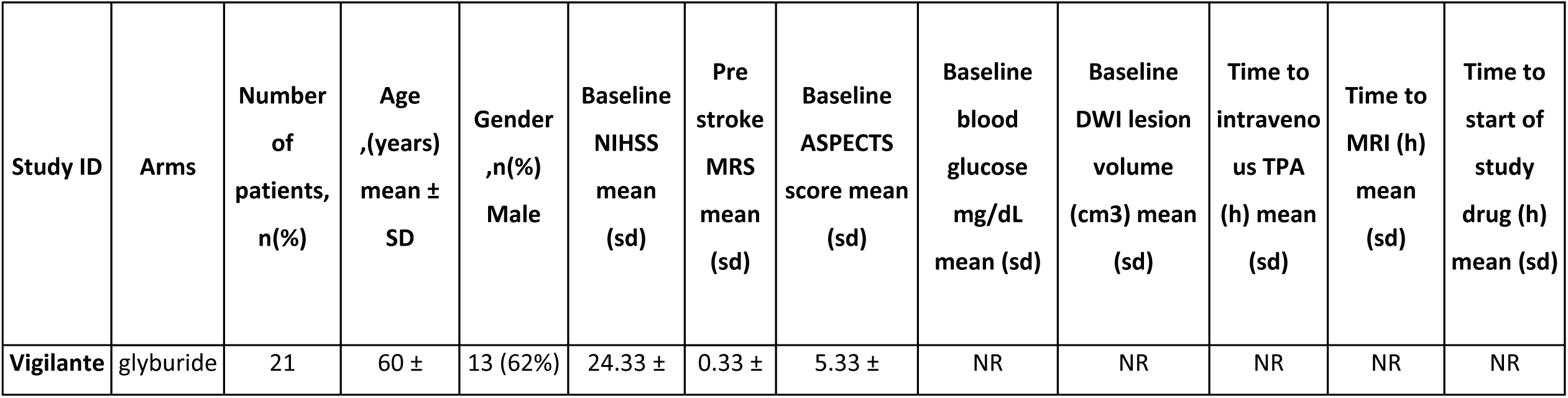
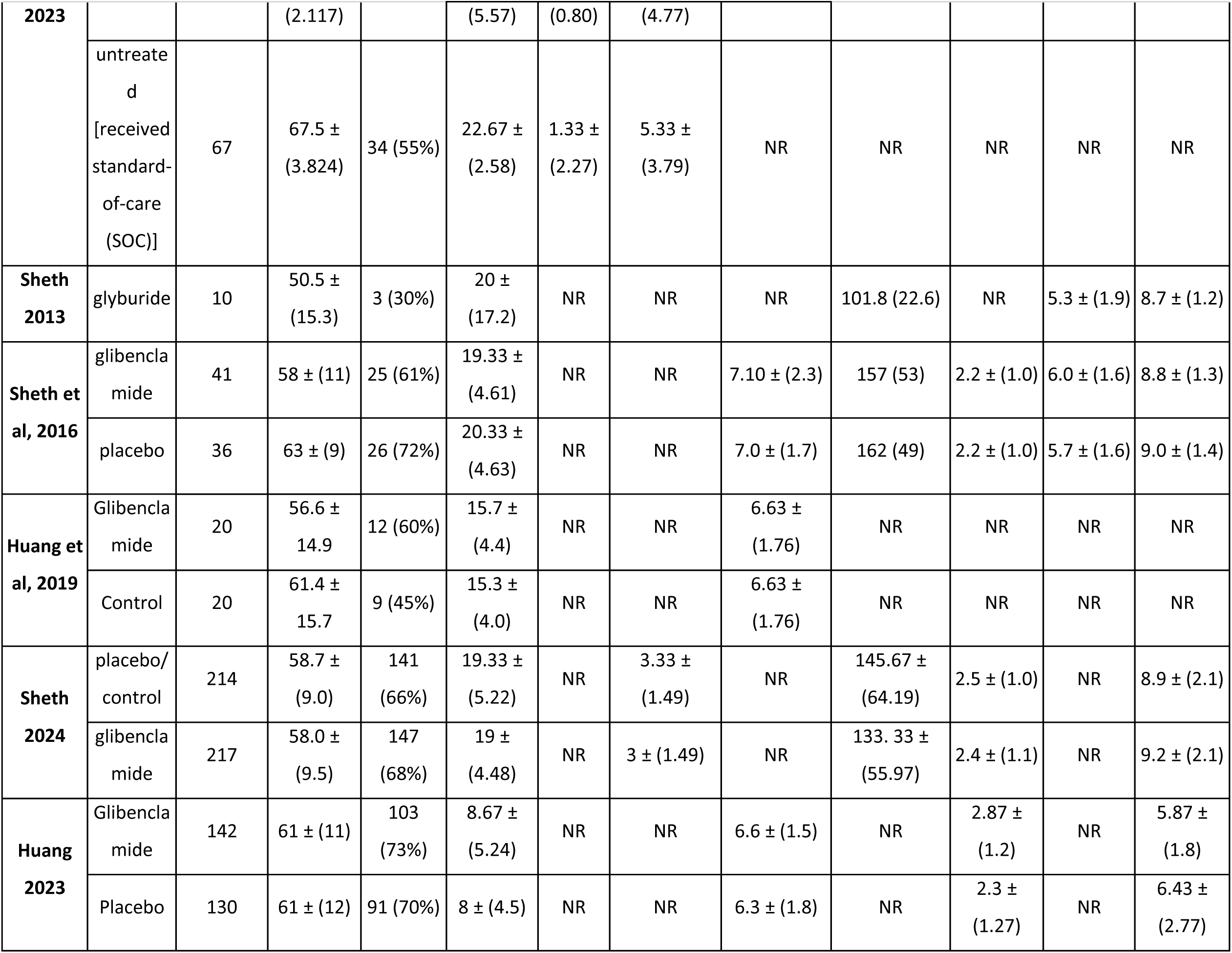
Table of Baseline Characteristics.

#### 3.2.1 Risk of Bias Assessment

The studies included in this meta-analysis show a range of methodological qualities. The risk of bias 2 tool showed that of the included four studies, one showed high risk, one showed some concerns, and two showed low risk. The study that showed the high risk of bias showed concern in its randomization process. One study, which showed some concern, had issues in missing outcome data and selection of reported results (**Figure 2**). Regarding the case control study, evaluation by the Newcastle-Ottawa Scale (NOS) tool showed it to have good quality based on three domains: selection → 4 stars, comparability → 2 stars, and exposure → 4 stars.

**Figure 2:**
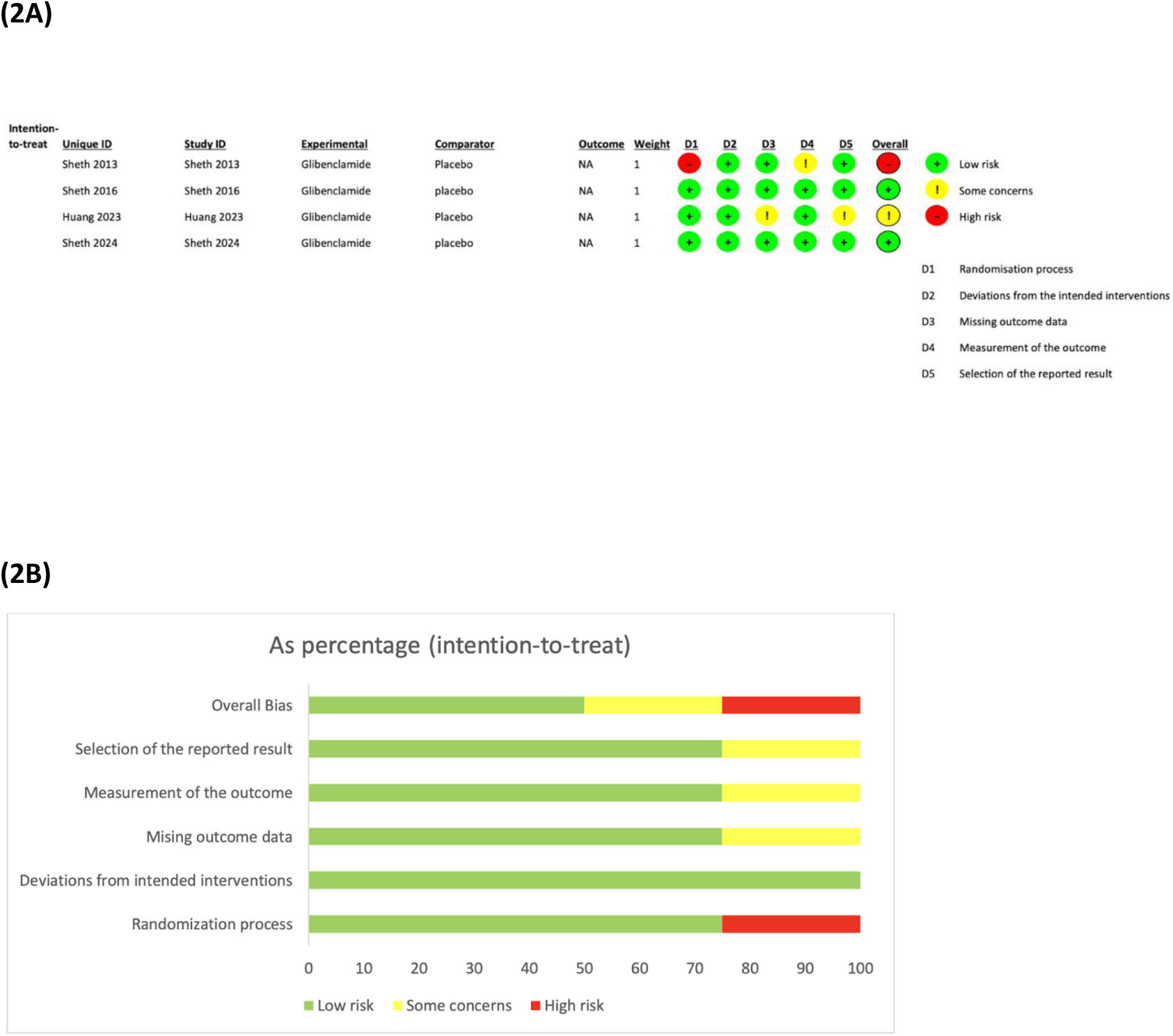
(A) Risk of Bias Tool Assessment and (B) Risk of Bias Tool Graph (2A).

Likewise, the cohort study showed good quality, with selection → 6 stars, comparability → 2 stars, and outcome → 4 stars.

### 3.3 Efficacy and Safety Outcomes

#### 3.3.1 Efficacy Outcomes

##### 3.3.1.1 Modified Rankin Score (mRS)

The Glibenclamide versus Placebo comparison shows that three studies reported on the Modified Rankin Score (mRS), which is a six-point scale used to measure the degree of disability or dependence post stroke, ranging from 0 (asymptomatic) to 6 (death). **Figure 3A** shows the overall effect of Glibenclamide versus Placebo on mRS (MD = 0.07, 95% CI: [−0.17, 0.31], P = 0.57). Analysis showed no heterogeneity (I^2^= 0%, P = 0.43).

**Figure 3:**
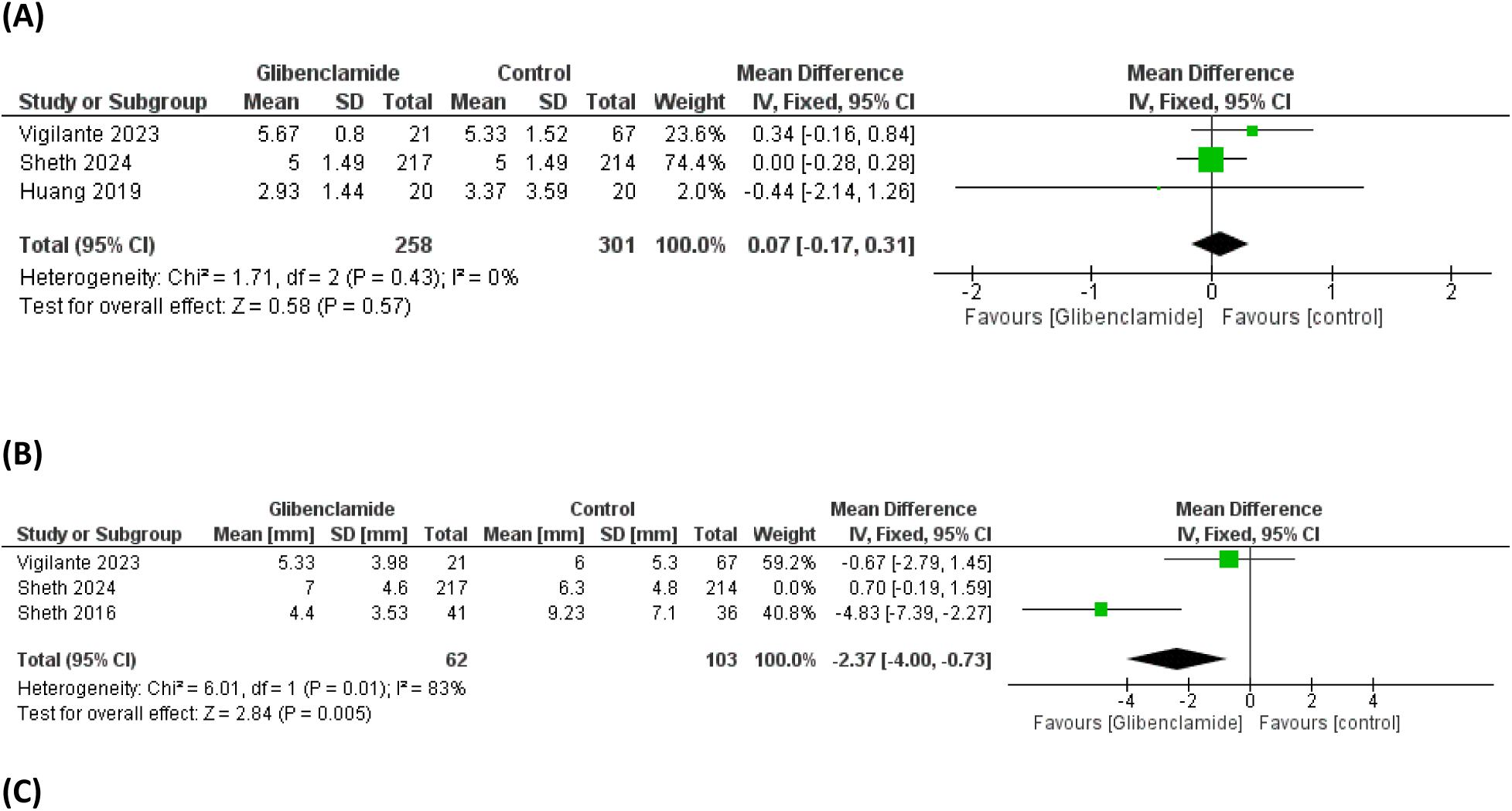
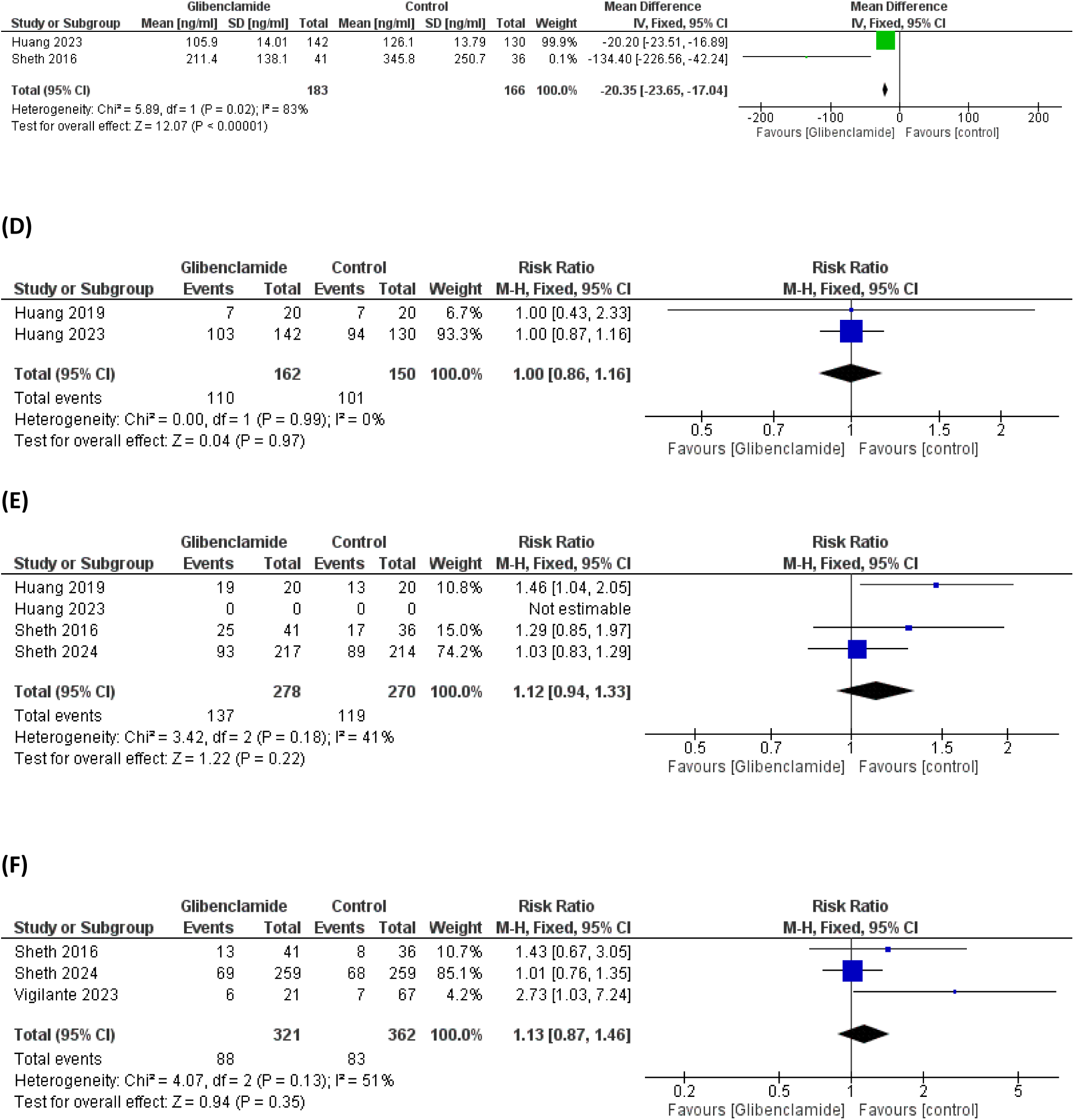
Forest plots show (A) Modified Rankin Score (B) Mean Midline Shift, (C) Mean MMP-9, (D) mRS of 0-2, (E) mRS of 0-4, and (F) Need for Decompressive Craniotomy.

##### 3.3.1.2 Mean Midline Shift at 72 hours in millimeters (mm)

Three studies reported on mean midline shift at 72 hours and showed a significant 2.37 millimeter decrease in midline shift with the use of Glibenclamide (MD = −2.37, 95% CI: [−4.00, - 0.73], P = 0.005). There was substantial heterogeneity found (I^2^= 83%, P = 0.01) that was not resolved by fixed analysis method or one study removal. (**Figure 3B**).

##### 3.3.1.3 Mean MMP-9 (ng/mL) during 24–72 Hours

Glibenclamide versus Placebo comparison shows that two studies reported on the mean MMP-9 during the 24-72 hour time period. **Figure 3C** shows a significant reduction in mean MMP-9 of 20.35 (MD = –20.35, 95% CI: [–23.65 to –17.04], P < 0.00001, I^2^= 0%). There was substantial heterogeneity found (I^2^= 83%, P = 0.02) that was not resolved by the fixed analysis method.

##### 3.3.1.4 mRS of 0-2

Two studies reported Modified Rankin scores of 0-2 when comparing Glibenclamide versus Placebo. **Figure 3D** shows the overall effect of (MD = 1.00, 95% CI: [0.86, 1.16], P = 0.97).

Analysis showed no heterogeneity (I^2^= 0%, P = 0.99).

##### 3.3.1.5 mRS of 0-4

The Glibenclamide versus Placebo comparison shows that four studies reported Modified Rankin scores of 0-4. **Figure 3E** shows the overall effect of Glibenclamide versus Placebo on mRS of 0-4 (MD = 1.12, 95% CI: [0.94, 1.33], P = 0.22). Analysis showed no significant heterogeneity (I^2^= 41%, P = 0.18).

##### 3.3.1.6 Need for Decompressive Craniotomy

The Glibenclamide versus Placebo comparison shows that three studies reported on the need for decompressive craniotomy (**Figure 3F**). The overall effect of (MD = 1.13, 95% CI: [0.87, 1.46], P = 0.35) was shown. No significant heterogeneity was found (I^2^= 51%, P = 0.13).

#### 3.3.2 Safety Outcomes

##### 3.3.2.1 All-Cause Mortality

Four studies reported on all-cause mortality when comparing Glibenclamide versus Placebo. **Figure 4A** shows the overall effect of Glibenclamide versus Placebo on mortality (MD = 1.07, 95% CI: [0.83, 1.38], P = 0.59). There was no significant heterogeneity shown (I^2^= 50%, P = 0.14).

**Figure 4:**
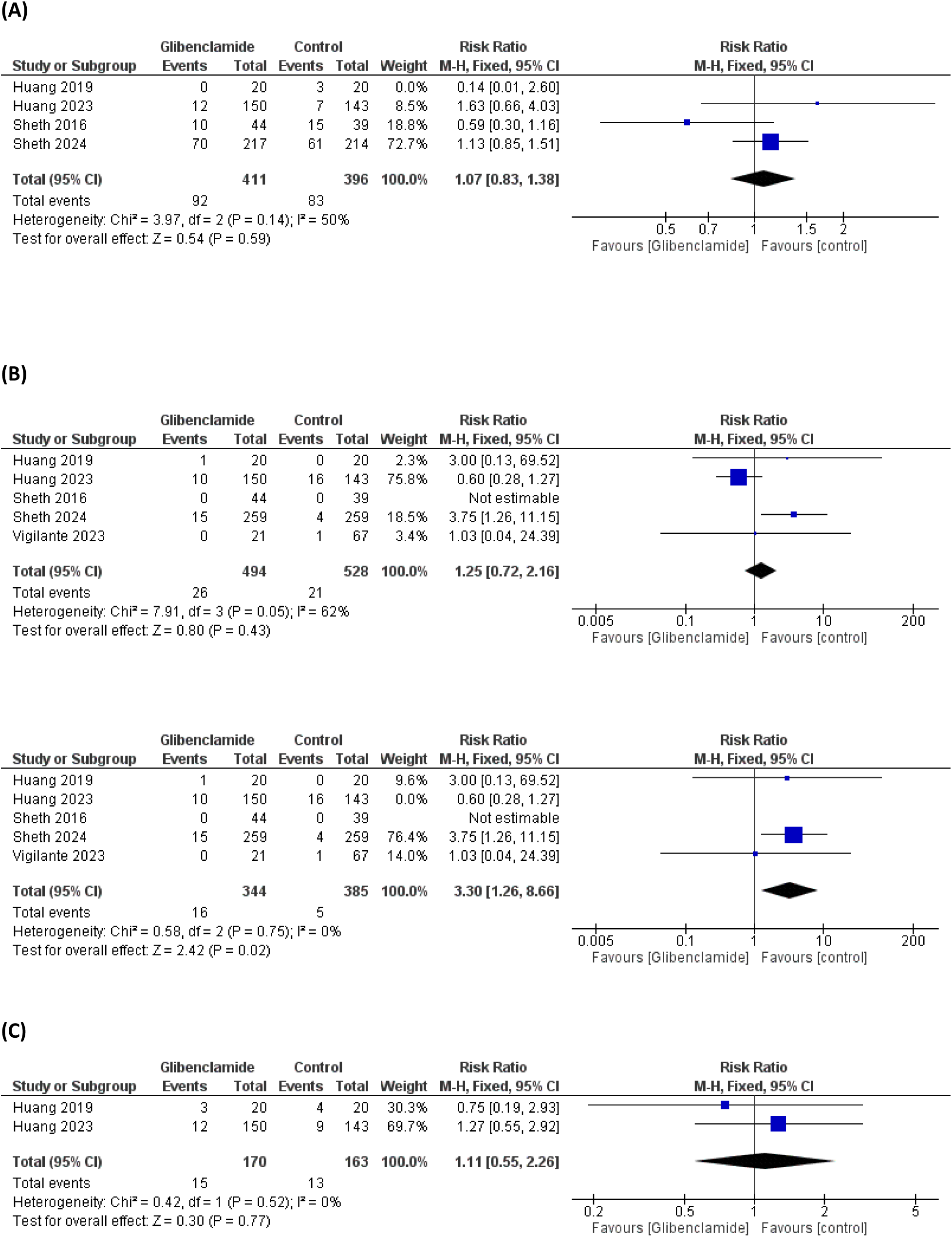
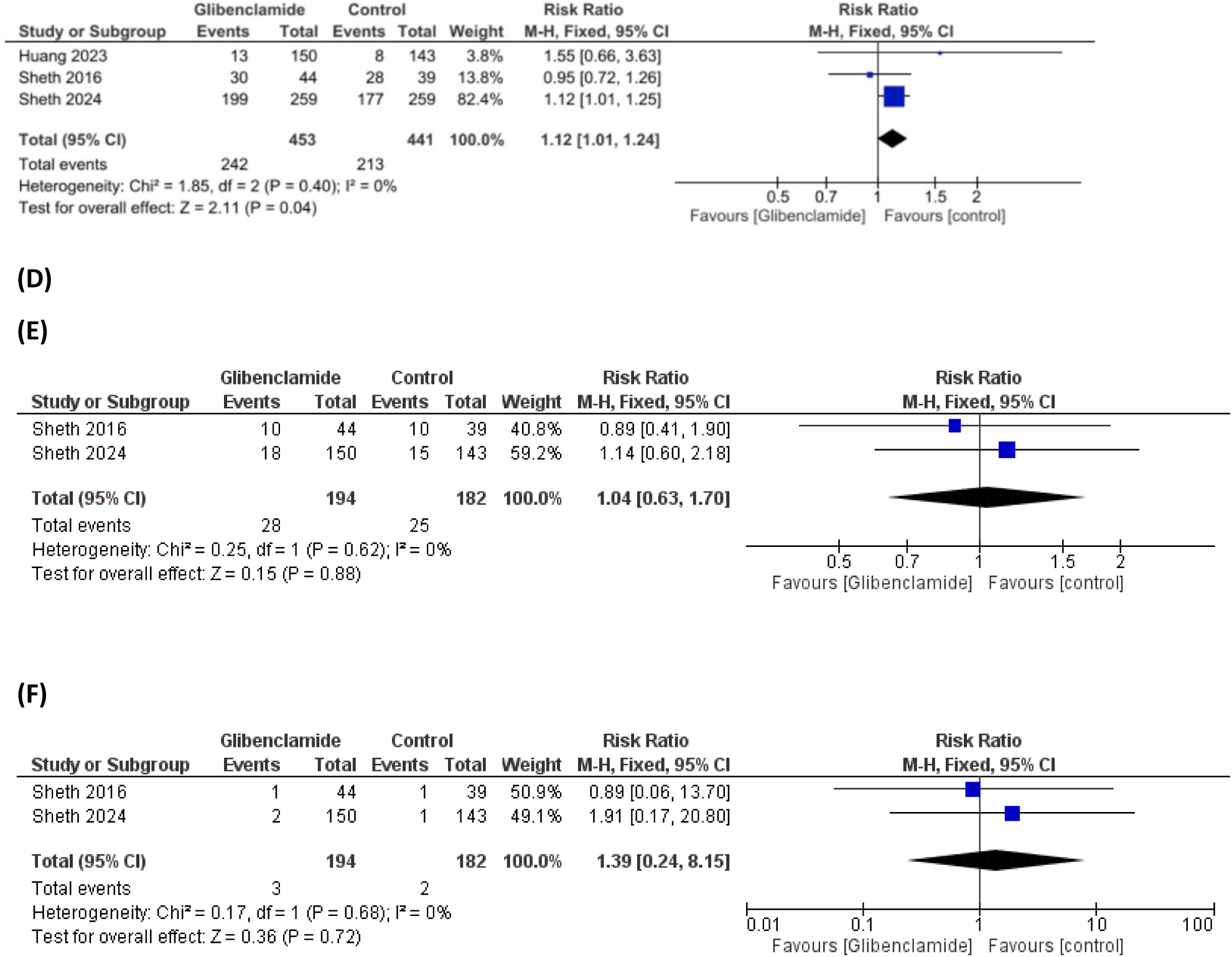
Forest plots show (A) All-Cause Mortality, (B) Hypoglycemia Events, (C) Neurological Deterioration, (D) Serious Adverse Events, (E) Cardiac Events, and (F) Cardiac Death.

##### 3.3.2.2 Hypoglycemia Events

Glibenclamide versus Placebo comparison shows that five studies reported on hypoglycemia events **(Figure 4B)**. The overall effect was not statistically significant (RR = 1.25, 95% CI: [0.72, 2.16], p = 0.43). There was substantial heterogeneity (I^2^ = 62%, p = 0.05).

Leaving out Sheth 2016 and comparing Glibenclamide versus placebo in four studies showed a higher risk of hypoglycemia with Glibenclamide use (RR = 3.30, 95% CI: [1.26, 8.66], p = 0.75). There was no significant heterogeneity shown (I^2^ = 0%, p = 0.02).

##### 3.3.2.3 Neurological Deterioration

Glibenclamide versus Placebo comparison shows that two studies reported on neurological deterioration events **(Figure 4C)**. The overall effect was not statistically significant (RR = 1.11, 95% CI: [0.55, 2.26], p = 0.77). There was no heterogeneity (I^2^ = 0%, p = 0.52).

##### 3.3.2.4 Serious Adverse Events

Glibenclamide versus Placebo comparison shows that three studies reported on serious adverse events **(Figure 4D)**. The overall effect was statistically significant, showing a 12% increased risk of developing a serious adverse event with Glibenclamide use (RR = 1.12, 95% CI: [1.01, 1.24], p = 0.04). There was no heterogeneity (I^2^ = 0%, p = 0.40).

##### 3.3.2.5 Cardiac Events

Glibenclamide versus Placebo comparison shows that two studies reported on cardiac events **(Figure 4E)**. The overall effect was not statistically significant (RR = 1.04, 95% CI: [0.63, 1.70], p = 0.88). There was no heterogeneity (I^2^ = 0%, p = 0.62).

##### 3.3.2.6 Cardiac Death

Glibenclamide versus Placebo comparison shows that two studies reported on cardiac death **(Figure 4F)**. The overall effect was not statistically significant (RR = 1.39, 95% CI: [0.24, 8.15], p = 0.72). There was no heterogeneity (I^2^ = 0%, p = 0.68).

## 4. Discussion

Acute ischemic stroke (AIS) remains a major global contributor to mortality and long-term disability (16). with malignant cerebral edema representing one of its most life-threatening complications (17). This meta-analysis evaluated the therapeutic efficacy and safety profile of Glibenclamide in AIS patients with cerebral edema.

Glibenclamide was originally developed as a sulfonylurea for diabetes management (18), and it has shown promise in limiting cerebral edema by inhibiting SUR1-TRPM4 channels implicated in edema formation (19). The findings demonstrate that Glibenclamide reduces key indicators of cerebral edema, including matrix metalloproteinase-9 (MMP-9) levels and midline shift, with no significant impact on the Modified Rankin Scale (mRS).

MMP-9 is a group of zinc-dependent endopeptidases linked to several neurological disorders, where neuroinflammation is a significant contributing factor (20). On the other hand, the brain midline shift (MLS) measures the extent of the mass effect caused by intracranial lesions like hematomas, brain tumors, strokes, and traumatic brain damage. Patients’ prognosis and whether they need emergent surgery can both be predicted by brain MLS (21). On the other hand, the Modified Rankin Scale is a scale that ranks overall impairment following a stroke. It is the most comprehensive and extensively used primary outcome measure in acute stroke studies (22). This scale, which ranks patients’ impairment from 0 (no symptoms) to 6 (death), represents the entire spectrum of functional states post-stroke and is seemingly simple (23).

### 4.1 Interpretation of Findings and Comparison with Existing Literature

This analysis shows that Glibenclamide is associated with reduced cerebral edema and inflammatory markers post-stroke, especially MMP-9 levels and midline shift. These findings support its potential role in altering early post-stroke pathophysiology. However, this review shows no significant observed improvements in functional outcomes such as modified Rankin Scale (mRS) scores or the need for decompressive craniectomy. This gap between the improvement in biomarkers and the lack of significant clinical benefit may be attributed to several factors, such as variability in stroke severity, timing of administration, and post-stroke care practices.

This suggests that Glibenclamide has a potential role in attenuating secondary brain injury. Given its likely benefit, its use was associated with a higher risk of hypoglycemia and other serious adverse events, underscoring the need for careful patient selection and monitoring Our findings of a statistically significant reduction in midline shift with Glibenclamide align with the systematic review by Wu et al. published in 2018 (24). While their review is focused on prediction rather than intervention, their conclusions highlight that Glibenclamide significantly reduces midline shift, which is a complication of the cerebral edema that happens post-stroke. Unlike our analysis, however, their review did not evaluate treatment-related adverse events like hypoglycemia or other adverse events. These side effects are of great importance, as they require close monitoring, particularly in populations at risk for glucose fluctuations.

Our findings are also largely consistent with a meta-analysis published by Fan et al in 2025 that evaluates Glibenclamide use in patients with severe hemispheric infarction (25). Fan et al study included only two studies, while our meta-analysis included six studies, including observational studies, in order to reach more comprehensive results. Like our results, the review reports no significant improvement in the modified Rankin Scale (mRS). Both reviews found an increased risk of hypoglycemia associated with glyburide, reinforcing the need for caution in its clinical application. Unlike their analysis, our analysis demonstrates a significant reduction in MMP-9 levels and midline shift, suggesting a potential systematic benefit of Glibenclamide in reducing inflammation and mass effect that was not captured in the above-mentioned analysis. These findings support further investigation into the biomarker and imaging-based effects of Glibenclamide beyond traditional clinical endpoints.

Another meta-analysis, the HERMES meta-analysis (26), did not observe a meaningful effect of thrombectomy or reperfusion on midline shift in patients with large hemispheric infarction. Interestingly, their exploratory subgroup analysis suggested that thrombectomy may even increase midline shift in patients with severe baseline imaging features. This highlights the complexity of edema evolution in the context of reperfusion. These findings are important, as they give potential for Glibenclamide to be used as a pharmacologic method to mitigate cerebral edema rather than an interventional approach, especially in patient populations where mechanical intervention alone may not suffice or may exacerbate swelling.

Comparing Glibenclamide with other antidiabetic agents, results seem to vary. According to a study done by Akhtar et al in 2022, diabetic stroke patients who received Metformin prior to treatment had significantly improved 90-day mRS outcomes and decreased mortality (27), indicating a functional benefit not seen with Glibenclamide. Metformin is an activator of the AMP-activated protein kinase (AMPK) (28), which is a sensor of cellular energy status that regulates cellular energy balance and glycolipid metabolism. In recent years, the anti-inflammatory function of AMPK and associated signaling cascades has gained widespread interest due to its impact on metabolic disorders (29). This provides greater support for the idea that Metformin may have effects on stroke recovery that go beyond glycemic management.

Another group of antidiabetic drugs that we compared our results to is Sodium-glucose cotransporter 2 (SGLT2) inhibitors. SGLT-2 inhibitors are a class of antidiabetic medications that work by reducing the kidneys’ reabsorption of glucose by acting on SGLT-2 receptors (30). A study by Yang et al published in 2024 showed that although stroke patients who were prescribed SGLT-2 inhibitors during admission had no significant differences in mRS scores upon discharge from the ones who did not, patients who received SGLT-2 inhibitors during admission had a significantly higher chance of having a better mRS score 3 months post stroke than those who did not (31).

While Glibenclamide shows promise in influencing the mechanisms that drive cerebral edema, its clinical application is still limited. The lack of long-term functional improvement highlights the multifactorial nature of AIS outcomes, which are influenced by more than just edema control. Future studies should focus on larger, well-designed randomized controlled trials (RCTs) to determine the most appropriate patient populations, dosing regimens, and treatment windows to assess the overall clinical benefit of Glibenclamide in this context.

### 4.2 Strengths and Limitations

This meta-analysis integrates data from both randomized controlled trials and high-quality observational studies, enhancing its relevance to real-world clinical settings. By examining both physiological and clinical endpoints, it offers a comprehensive assessment of Glibenclamide’s therapeutic potential.

Nonetheless, several limitations must be acknowledged. The small number of included studies (n=6), with only five contributing to pooled analyses, limits statistical power and generalizability. Heterogeneity in study design, patient populations, and intervention protocols may have influenced outcome estimates, despite efforts to reduce bias through standardized methodology and sensitivity analyses. Significant heterogeneity was observed in some outcomes, such as midline shift (I² = 83%), potentially due to differences in assessment methods or inclusion criteria. Additionally, short follow-up durations in most studies precluded evaluation of long-term outcomes like cognitive function or stroke recurrence. Finally, potential publication bias and language restrictions may have led to the omission of relevant studies, particularly non-English publications.

### 4.3 Implications for Clinical Practice

While current evidence does not support the routine use of Glibenclamide for improving functional outcomes after AIS, its potential role in controlling cerebral edema and inflammatory markers is promising. In selected high-risk patients, such as those with large infarcts or early signs of malignant edema, Glibenclamide could serve as an adjunctive therapy to reduce the need for invasive interventions. However, clinicians must remain cautious about the drug’s side effects, especially hypoglycemia, and ensure proper monitoring and patient selection.

Our findings underscore the complexity of stroke pathophysiology and highlight the need for therapies that address both early edema formation and long-term recovery. While Glibenclamide may be beneficial in managing the former, its effects on the latter are still uncertain.

### 4.4 Future Directions

Large, multicenter RCTs with consistent procedures and well-defined inclusion criteria need to be given priority in future research. Subgroups that benefit most from Glibenclamide may be found by stratifying individuals according to baseline edema severity and infarct features.

Important areas for additional research include assessing combination therapy with other neuroprotective drugs, optimizing dose schedules, and determining the best time to administer medications. Moreover, the use of biomarkers like MMP-9 as early surrogate endpoints should improve the design and sensitivity of future trials. Long-term outcomes, such as neurocognitive function, disability-adjusted life years (DALYs), and caregiver burden, should also be evaluated to identify the overall therapeutic impact.

## 5. Conclusion

This meta-analysis summarizes the available information on Glibenclamide in AIS complicated by cerebral edema. While the medicine can diminish edema-related biomarkers such as MMP-9 and midline shift, it does not appear to improve functional results or decrease the requirement for decompressive surgery. Additionally, its use is linked to an increased risk of hypoglycemia and significant adverse effects.

Although Glibenclamide may influence early edema-related processes, the lack of persistent clinical benefit implies that stroke recovery is dependent on variables other than edema control. Its role in AIS care is unknown, and it should be limited to research settings or tightly monitored clinical use until more evidence is available.

## Declarations

### Author contribution

AK, AT, and ME: conceptualization and methodology.

AR, AT, and BA: investigation and data curation, screening, and data extraction.

AK: formal analysis.

SO, NM, BA, and AR: Writing (Original Draft).

ME: Supervision.

ME: Project administration.

ME, BA, AK, SO, and NM: Writing - Review & Editing.

All authors read and approved the final content.

## Ethics approval

Not applicable

## Consent to participate

Not applicable.

## Consent for publication

Not applicable.

## Availability of data and material

All data generated or analyzed during this study are included in this published article or the data repositories listed in References.

## Code availability

Not applicable.

## Acknowledgment

Not applicable.

## References

1. Grysiewicz RA, Thomas K, Pandey DK. Epidemiology of Ischemic and Hemorrhagic Stroke: Incidence, Prevalence, Mortality, and Risk Factors. Neurol Clin. 2008 Nov;26(4):871–95.

2. Donkor ES. Stroke in the 2 1 s t Century: A Snapshot of the Burden, Epidemiology, and Quality of Life. Stroke Res Treat. 2018 Nov 27;2018:1–10.

3. Khanna A, Walcott BP, Kahle KT, Simard JM. Effect of glibenclamide on the prevention of secondary brain injury following ischemic stroke in humans. Neurosurg Focus. 2014 Jan;36(1):E11.

4. Gu Y, Zhou C, Piao Z, Yuan H, Jiang H, Wei H, et al. Cerebral edema after ischemic stroke: Pathophysiology and underlying mechanisms. Front Neurosci. 2022 Aug 18;16:988283.

5. Cook AM, Morgan Jones G, Hawryluk GWJ, Mailloux P, McLaughlin D, Papangelou A, et al. Guidelines for the Acute Treatment of Cerebral Edema in Neurocritical Care Patients. Neurocrit Care. 2020 Jun;32(3):647–66.

6. Luzi L, Pozza G. Glibenclamide: an old drug with a novel mechanism of action? Acta Diabetol. 1997 Dec 15;34(4):239–44.

7. Sheth KN, Albers GW, Saver JL, Campbell BCV, Molyneaux BJ, Hinson HE, et al. Intravenous glibenclamide for cerebral oedema after large hemispheric stroke (CHARM): a phase 3, double-blind, placebo-controlled, randomised trial. Lancet Neurol. 2024 Dec;23(12):1205–13.

8. Page MJ, McKenzie JE, Bossuyt PM, Boutron I, Hoffmann TC, Mulrow CD, et al. The PRISMA 2020 statement: an updated guideline for reporting systematic reviews. Syst Rev. 2021 Dec;10(1):89.

9. Higgins JPT, Thomas J, Chandler J, Cumpston M, Li T, Page MJ, Welch VA (editors). Cochrane Handbook for Systematic Reviews of Interventions version 6.5 (updated August 2024). Cochrane, 2024. Available from www.training.cochrane.org/handbook.

10. Ouzzani M, Hammady H, Fedorowicz Z, Elmagarmid A. Rayyan—a web and mobile app for systematic reviews. Syst Rev. 2016 Dec;5(1):210.

11. Sterne JAC, Savović J, Page MJ, Elbers RG, Blencowe NS, Boutron I, et al. RoB 2: a revised tool for assessing risk of bias in randomised trials. BMJ. 2019 Aug 28;l4898.

12. Stang A. Critical evaluation of the Newcastle-Ottawa scale for the assessment of the quality of nonrandomized studies in meta-analyses. Eur J Epidemiol. 2010 Sep;25(9):603–5.

13. Wan X, Wang W, Liu J, Tong T. Estimating the sample mean and standard deviation from the sample size, median, range and/or interquartile range. BMC Med Res Methodol. 2014 Dec;14(1):135.

14. Altman DG, Bland JM. Standard deviations and standard errors. BMJ. 2005 Oct 15;331(7521):903.

15. Higgins JPT, Thomas J, Chandler J, Cumpston M, Li T, Page MJ, Welch VA. Cochrane Handbook for Systematic Reviews of Interventions version 6.3 (updated February 2022). Cochrane, 2022 [Internet]. Higgins JPT, Thomas J, Chandler J, Cumpston M, Li T, Page MJ, Welch VA, editor. [cited 2023 May 7]. Available from: https://training.cochrane.org/handbook.

16. Lui F, Hui C, Khan Suheb MZ, et al. Ischemic Stroke. [Updated 2025 Feb 21]. In: StatPearls [Internet]. Treasure Island (FL): StatPearls Publishing; 2025 Jan-. Available from: https://www.ncbi.nlm.nih.gov/books/NBK499997/.

17. Foroushani HM, Hamzehloo A, Kumar A, Chen Y, Heitsch L, Slowik A, et al. Accelerating Prediction of Malignant Cerebral Edema After Ischemic Stroke with Automated Image Analysis and Explainable Neural Networks. Neurocrit Care. 2022 Apr;36(2):471–82.

18. Sola D, Rossi L, Schianca GPC, Maffioli P, Bigliocca M, Mella R, et al. State of the art paper Sulfonylureas and their use in clinical practice. Arch Med Sci. 2015;4:840–8.

19. Jha RM, Rani A, Desai SM, Raikwar S, Mihaljevic S, Munoz-Casabella A, et al. Sulfonylurea Receptor 1 in Central Nervous System Injury: An Updated Review. Int J Mol Sci. 2021 Nov 2;22(21):11899.

20. Gasche Y. Matrix metalloproteinases and diseases of the central nervous system with a special emphasis on ischemic brain. Front Biosci. 2006;11(1):1289.

21. Wu AR, Hsieh SY, Chou HH, Lai CS, Hung JY, Wang B, et al. Deep learning-based prediction of mortality using brain midline shift and clinical information. Heliyon. 2025 Jan;11(2):e41271.

22. Saver JL, Filip B, Hamilton S, Yanes A, Craig S, Cho M, et al. Improving the Reliability of Stroke Disability Grading in Clinical Trials and Clinical Practice: The Rankin Focused Assessment (RFA). Stroke. 2010 May;41(5):992–5.

23. Pożarowszczyk N, Kurkowska-Jastrzębska I, Sarzyńska-Długosz I, Nowak M, Karliński M. Reliability of the modified Rankin Scale in clinical practice of stroke units and rehabilitation wards. Front Neurol. 2023 Mar 3;14:1064642.

24. Wu S, Yuan R, Wang Y, Wei C, Zhang S, Yang X, et al. Early Prediction of Malignant Brain Edema After Ischemic Stroke: A Systematic Review and Meta-Analysis. Stroke. 2018 Dec;49(12):2918–27.

25. Fan L, Xu J, Wang T, Yang K, Bai X, Yang W. Sulfonylurea drugs for people with severe hemispheric ischemic stroke. Cochrane Central Editorial Service, editor. Cochrane Database Syst Rev [Internet]. 2025 Mar 11 [cited 2025 May 11];2025(3). Available from: http://doi.wiley.com/10.1002/14651858.CD014802.pub2

26. Ng FC, Yassi N, Sharma G, Brown SB, Goyal M, Majoie CBLM, et al. Cerebral Edema in Patients With Large Hemispheric Infarct Undergoing Reperfusion Treatment: A HERMES Meta-Analysis. Stroke. 2021 Nov;52(11):3450–8.

27. Akhtar N, Singh R, Kamran S, Babu B, Sivasankaran S, Joseph S, et al. Diabetes: Chronic Metformin Treatment and Outcome Following Acute Stroke. Front Neurol. 2022 Apr 26;13:849607.

28. Venna VR, Li J, Hammond MD, Mancini NS, McCullough LD. Chronic metformin treatment improves post-stroke angiogenesis and recovery after experimental stroke. Eur J Neurosci. 2014 Jun;39(12):2129–38.

29. Xu Y, Bai L, Yang X, Huang J, Wang J, Wu X, et al. Recent advances in anti-inflammation via AMPK activation. Heliyon. 2024 Jul;10(13):e33670.

30. Jasleen B, Vishal GK, Sameera M, Fahad M, Brendan O, Deion S, et al. Sodium-Glucose Cotransporter 2 (SGLT2) Inhibitors: Benefits Versus Risk. Cureus [Internet]. 2023 Jan 18 [cited 2025 May 11]; Available from: https://www.cureus.com/articles/132075-sodium-glucose-cotransporter-2-sglt2-inhibitors-benefits-versus-risk

31. Yang W, Kim JM, Chung M, Ha J, Kang DW, Lee EJ, et al. Sodium-Glucose Cotransporter 2 Inhibitor Improves Neurological Outcomes in Diabetic Patients With Acute Ischemic Stroke. J Stroke. 2024 May 31;26(2):342–6.

